# Advancing Woman-Centered Global Health Simulations: Improved Accuracy Through Detailed Demographic Modelling in FPsim

**DOI:** 10.1101/2025.09.17.25336008

**Authors:** Emily Driano, Paula Sanz-Leon, Robyn Stuart, Michelle O’Brien, Marita Zimmermann

## Abstract

**Background:** FPsim is an agent-based model designed to simulate individual family planning (FP) behaviors and outcomes, capturing biological and behavioral heterogeneity among women. The initial release of the model enabled users to estimate the impact of new contraceptive technologies and interventions, but it lacked key contextual factors—particularly those linked to women’s socioeconomic status—that influence contraceptive dynamics. In this work, we enhanced FPsim by incorporating additional demographic attributes, including education level, wealth quintile, and urban/rural residence. These attributes influence, and are influenced by, contraceptive behaviors, allowing for a more nuanced simulation of family planning outcomes.

**Methods:** The key enhancements to the FPsim model are/were: (1) refining the behavioral algorithms that determine a woman’s probability of contraceptive uptake, method choice, and—importantly—the duration a woman remains on a given method, and (2) integrating additional social and demographic factors to better reflect real-world contraceptive choice. We added demographic attributes of education, urban/rural residence, and wealth quintile. By capturing how social context influences duration and switching behavior, FPsim v2.0 better reflects real-world contraceptive trajectories and heterogeneity across population subgroups.

**Contribution:** FPsim v2.0 advances a more comprehensive and person-centered approach to modeling FP behaviors. The new model embeds additional social and demographic diversity into individual agent profiles, offering the ability to simulate more tailored and effective interventions. Further validation in diverse settings is necessary to generalize these findings and maximize the model’s utility.

## Background

Increasingly, researchers and policymakers in family planning (FP) have attempted to broaden the goals of family planning programs beyond health outcomes. For example, FP offers young girls and adolescent women the ability to avoid pregnancy during school years, and to achieve their educational goals. Additional understanding of the social and demographic drivers of FP and its consequences can give us greater insight into how global health programs can better meet women’s needs.

In this manuscript, we present the methodology we use, including both statistical modeling and agent-based model parameterization, to extend the family planning simulator (FPsim) as a step toward more granular modeling for global health programming. FPsim is an agent-based model designed to simulate individual family planning (FP) behaviors and outcomes(13). This model incorporates biological and behavioral heterogeneity among women, capturing life events such as contraceptive use, pregnancy, and fertility outcomes. Leveraging demographic data, FPsim provides a detailed understanding of FP dynamics at the individual and aggregate levels, offering valuable insights into the potential effectiveness of interventions, such as introducing new contraceptive technologies. By simulating diverse and realistic individual trajectories, FPsim aids researchers and policymakers in understanding and improving family planning strategies on both local and global scales. However, the initial release of FPsim had a key limitation in that the outcomes of family planning were limited to mCPR and maternal health impacts. This manuscript describes the enhancements we introduced to resolve this limitation by incorporating the broader impacts of family planning on her life.

This work was undertaken as a part of the Family Planning Impact Consortium (FP-Impact), which aims to conduct a research, modeling and dissemination project centered on how support for women’s family planning intentions provides a path to women’s empowerment and economic development. In this paper, we describe the methodology used to implement FPsim v2.0, an improved version of the agent-based model which includes improved estimation of contraceptive use. We used a calibration of the model to a Kenya-like setting for this analysis.

## Methods

### Model structure

FPsim is an open-source agent-based model that can be calibrated for various contexts, with initial implementations in Senegal and ongoing development for Kenya and Ethiopia^1^. Agent-based models (ABMs) are rule-based computational models, in which researchers can simulate cohorts of individual agents and run hypothetical simulated scenarios. Agents are faced with decisions and events throughout their life course, most of which are decided probabilistically. In FPsim, each woman begins with her own individual underlying biological fecundity. They can then experience sexual debut, sexual activity, contraceptive choice, conception, abortion, miscarriage, stillbirth, a live birth, infant mortality, and/or maternal mortality. Agents encounter these events with different probabilities based on data from the context, and for most events, broken down into age and time period. For instance, the probability of experiencing infant mortality will be higher in high mortality areas, will decline over time as we see health systems improve in the context, and will be relatively higher for our youngest mothers(13).

In brief, the core components of the model – fecundability and contraceptive choice, are represented individually and probabilistically, following age-specific patterns observed in demographic data and prospective cohort studies. Each agent goes through a series of individualized, data-driven, probabilistic decisions each time step related to pregnancy and contraceptive choice (Figure 1).

**Figure 1.**
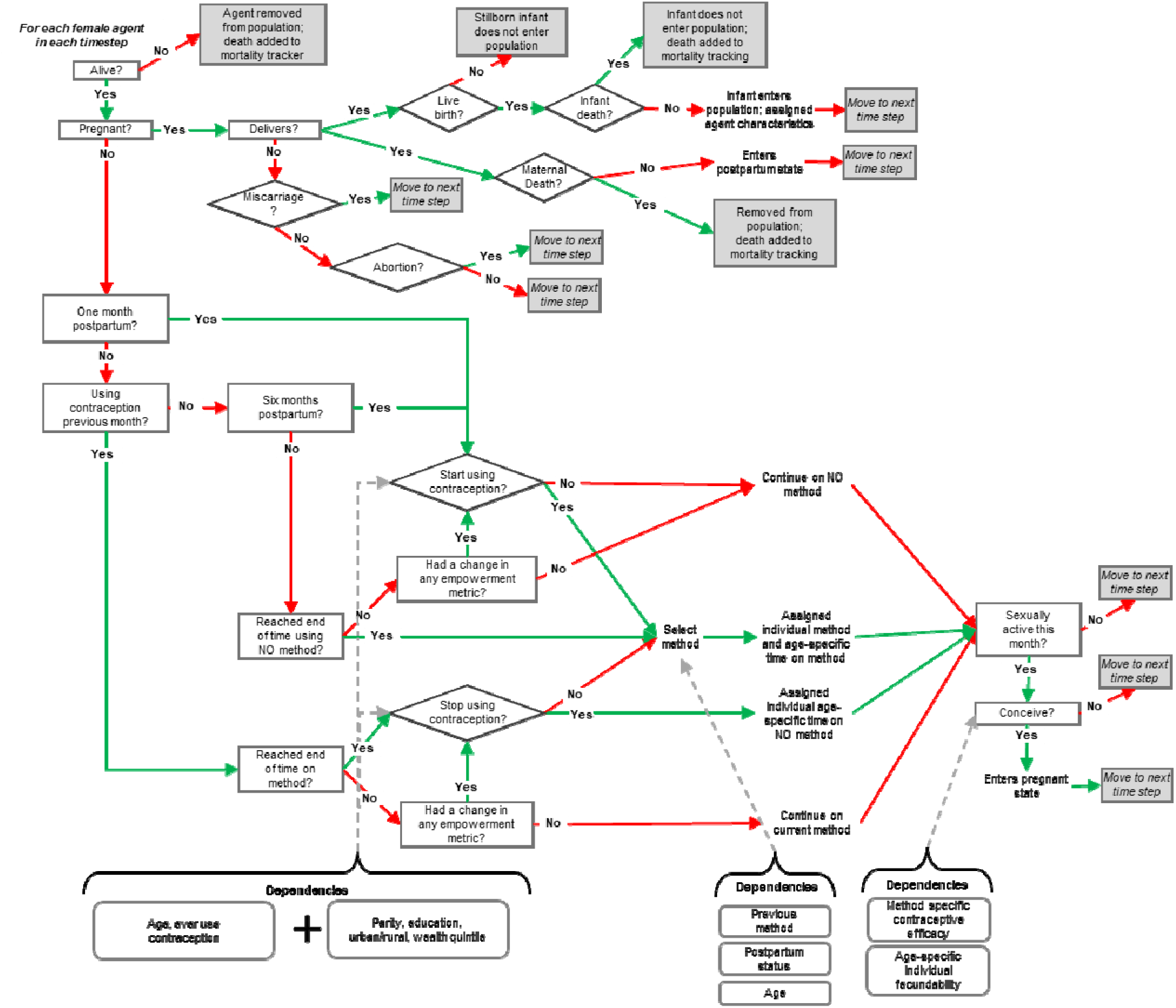
Decision map displaying contraception and pregnancy-related events in FPsim. Reading from the top, each agent in each timestep is checked for live status, and if alive, is checked for pregnancy status. If pregnant, the agent enters the pregnancy and childbirth module. If not, she continues on by checking if she is postpartum and if she was previously using contraception. She then walks through a series of decisions to determine if she will start or stop using contraception. If she is sexually active, she is eligible for conception.

### Data source

FPsim v2.0 leverages multiple sources of data for calibration. The key data source for this analysis and for the enhancements described in this study is the Kenya Performance Monitoring for Action (PMA) data(14). PMA is a nationally representative survey completed annually. Survey participants are women aged 15 to 49 who are asked about their background, birth histories, family planning methods, fertility preferences, community norms, aspirations, intentions, empowerment, and more. The survey is based on a multi-stage cluster design with urban-rural counties as strata. Households are randomly sampled within each representative geographical cluster. Beginning in 2019, PMA implemented a panel design embedded within the cross-sectional surveys, allowing for longitudinal analysis. We used three years of longitudinal data from 2019, 2020, and 2021. Where we used data from two timepoints, as described in the functions below, we combined data from 2019 to 2020 and 2020 to 2021.

### Demographic attributes added to FPsim v2.0

To implement the model improvements described below, we added three new demographic attributes to FPsim v2.0: level of education, place of residence (urban/rural), and wealth quintile. All three attributes were initialized based on the distribution by age in the 2022 Kenya Demographic and Health Survey (DHS) for ages 15-49. Wealth quintile and residential setting were constant over the lifetime, while the level of education progressed over time.

Upon entering the model, each woman is assigned an education goal based on the distribution of years of education obtained by women over age 20 (assumed to have completed education) in the 2022 Kenya DHS, stratified by place of residence. Starting at age 6, she continues gaining one year of education each year until she reaches her objective, unless she has an interruption. This can either be a temporary disruption (interruption) due to pregnancy, or a permanent disruption (dropout) based on DHS age-parity dropout probabilities.

We assumed all women still in school would have their education interrupted during pregnancy. The probability of dropping out was based on Kenya PMA data from 2019-2021. This was calculated by finding the percentage of all women who had a pregnancy while still in school who stopped schooling within a year of the birth. This percentage was stratified by age and parity (1 vs 2+). For those who do not drop out, the mechanism to resume education is tied to postpartum duration. If a woman interrupts her education due to pregnancy, and has not dropped out, she will go back to school 9 months after delivery (half the 6-month postpartum duration). While qualitative and case-study research on the burden of adolescent pregnancy in Kenya is compelling(15), we do not yet have data on this mechanism of return, or the duration of absence. This approximation was instead informed by the re-entry criterion for learners aged < 18 from Kenya’s National School Re-entry Guidelines that pregnant learners shall re-enter school six months after delivery, and at the beginning of the next calendar year.

### Contraceptive use

We implemented contraceptive use as a three-step process in the model: decide if a woman is using contraception, 2) if so, which method she will use, and 3) for that method, how long she will use it.

#### Probability of using contraception

One of the most important aspects of the model is determining if a woman is using contraception. We implemented two different versions of a function for determining her probability of using contraception at that time, each with increasing levels of complexity. Each function is a survey-weighted GLMs with a quasi-binomial family. The model user can select which of the functions to use based on the data available and the research question.

The probability of using contraception is first assessed at the time of her sexual debut. It is then reassessed if she has reached the end of her time on a method (described in further detail below). Additionally, the probability of contraception is assessed at 1 month postpartum, and, if she does not start using a method then, again at 6 months postpartum. These postpartum functions have the same form as the non-postpartum functions but are parameterized using data from women at the time postpartum. Therefore, for each of the functions described below, three sets of coefficients are produced and implemented as FPsim v2.0 model inputs, 1-month postpartum, 6-months postpartum, and not postpartum.

1. Simple choice function (eq. 1) The covariate for this function was age group (15-18, 19-20, 21-25, 26-35, and 36+). We parameterized this function using PMA data, but it could be calculated using more widely available datasets such as the DHS for future model calibrations.

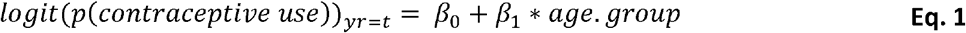
2. Detailed choice function (eq. 2) The covariates for this function were:
  ⍰ Age, implemented continuously with a cubic spline with knots at age 25 and 0
  ⍰ Previous contraceptive use, with an interaction with age
  ⍰ Parity, implemented as number of live births
  ⍰ Education, implemented as highest grade completed
  ⍰ Urban/rural
  ⍰ Wealth quintile
  We parameterized this function using PMA data, but it could be calculated using more widely available datasets such as the demographic and Health Survey (DHS) for future model calibrations.

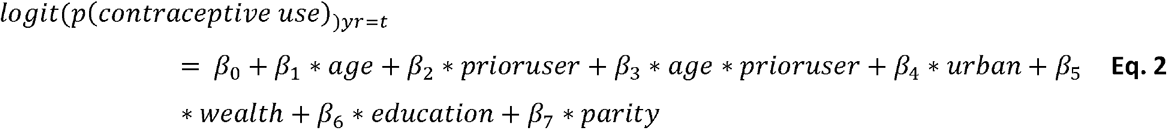

Resulting coefficient estimates for each of these modules can be found in the Appendix. We evaluated fit of the models from each function model using a calibration plot (Figure 2). Observed proportions of women using contraception were compared to predicted probabilities for contraceptive use. Models indicated good agreement between predicted and actual outcomes.

**Figure 2.**
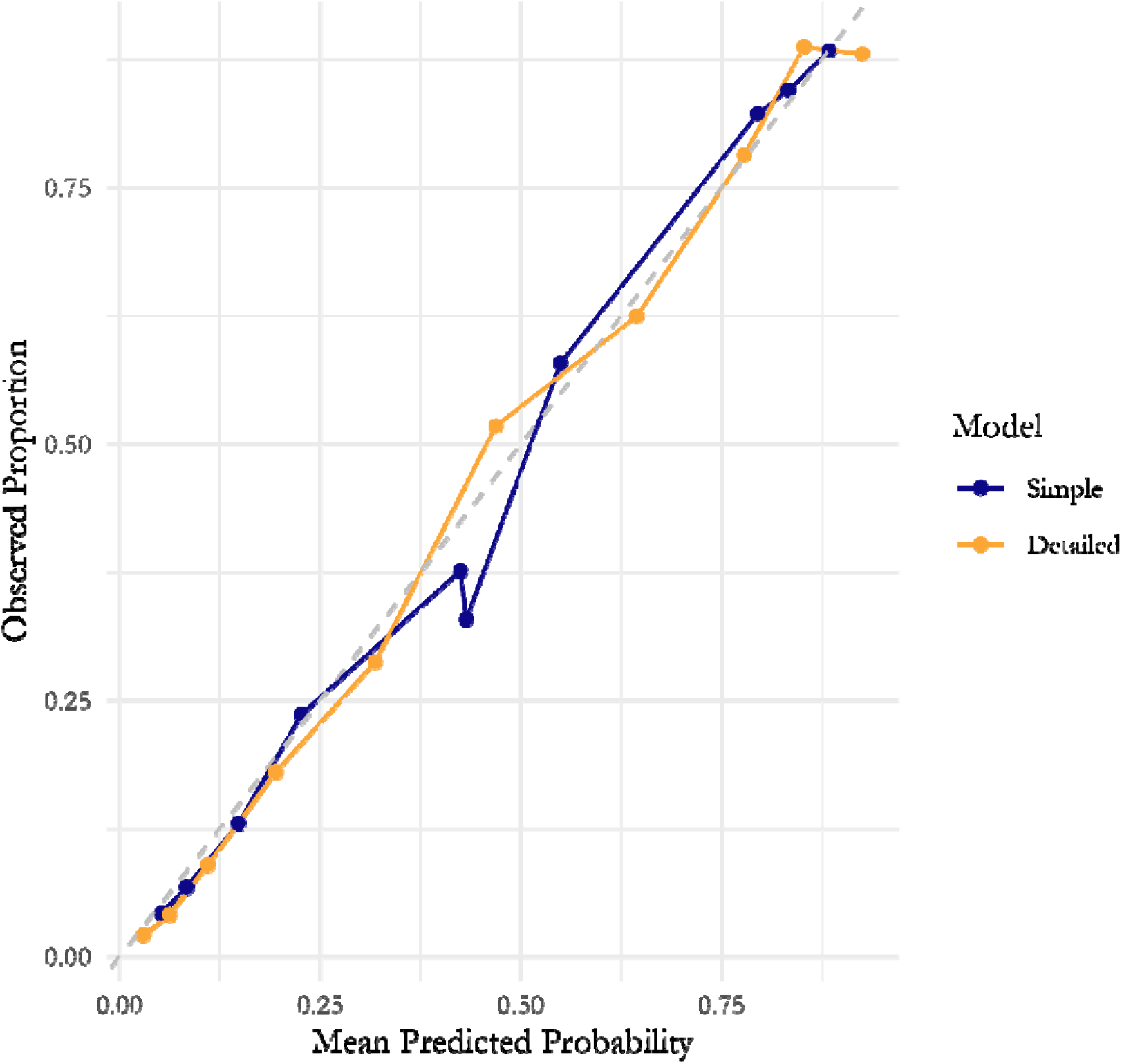
Calibration plots for simple logistic regression models across postpartum time subsets. Predicted probabilities compared to observed proportions for two models predicting current contraceptive use among women who are not currently postpartum: a simple predictive model using only age and prior contraceptive use (blue), and a more detailed demographic prediction model (yellow). Predicted probabilities were grouped into deciles, and for each bin, the mean predicted probability is plotted against the observed frequency of contraceptive use. The dashed diagonal line represents perfect calibration.

We adjust the probability of using contraception based on a time trend to account for population level changes in social norms and policies that we cannot capture in this individual model. To do this, we add a covariate to each of the functions for the time trend value * year / base year, where the time trend value is calculated as the average linear change in the contraceptive prevalence rate in Kenya from the base year.

#### Method choice

If a woman does begin to use contraception, she selects which method to use based on data from the contraceptive calendar in the 2022 Kenya DHS. The contraceptive calendar tracks which contraceptive method a woman is using, if any, for each month for up to 5 years prior to the survey. The calendar data was reformatted to represent which method a woman was using in each month, stratified by age group and which method she was using in the previous month. For 1-month postpartum, 6-months postpartum, and not postpartum, we produced age and previous method stratified method mix probabilities, which are used to select the method (Figure 3). While we would ideally stratify by further demographic factors, the sample size is not large enough to allow for further stratification.

**Figure 3.**
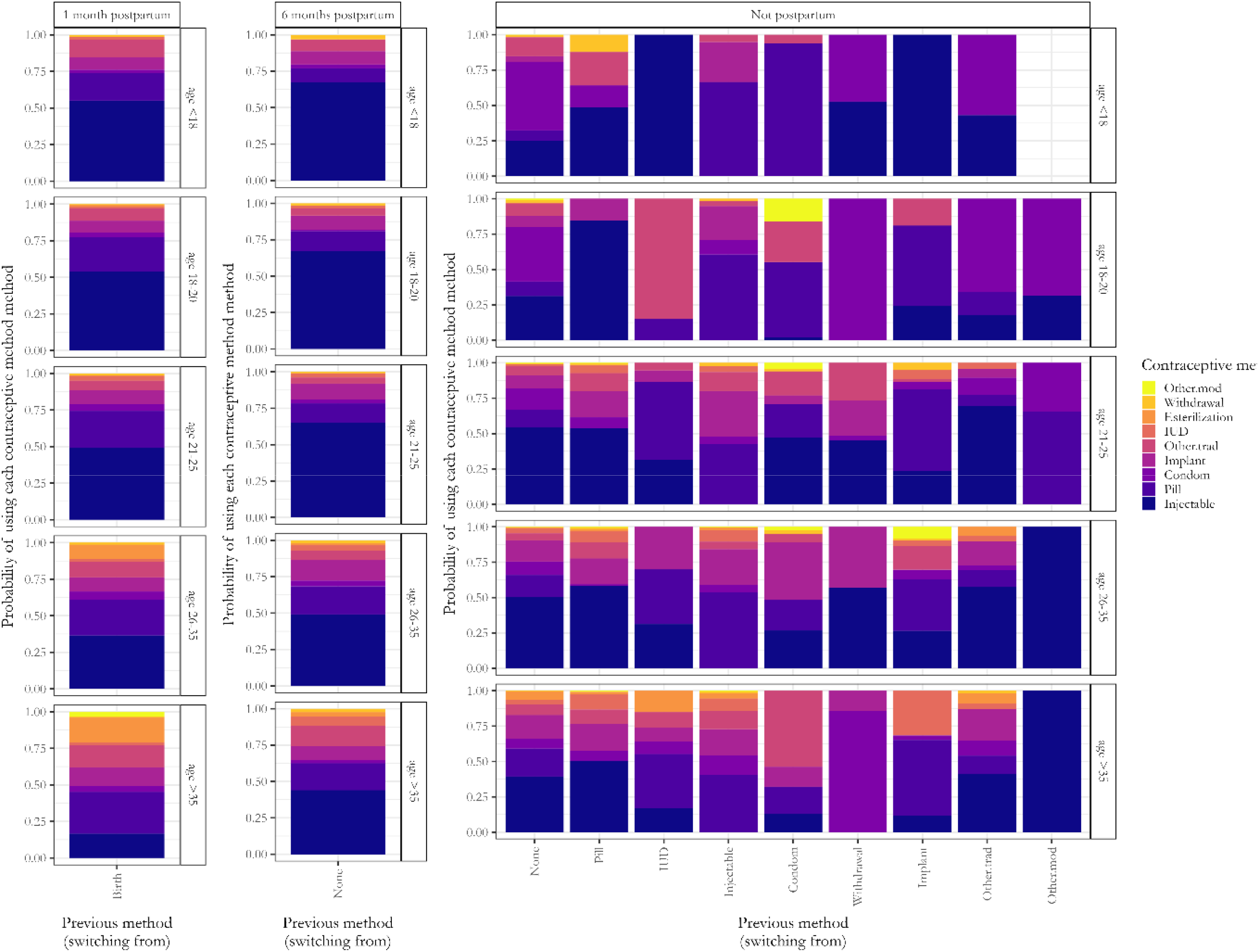
Method selection. When a woman is eligible chooses to start using contraception based on her individualized probability of using a method, she selects the specific method based on these percentages from the Kenya 2022 DHS, stratified by age group and which method she is switching from.

#### Time on method

Once a method is selected, or a woman decides not to use a method, she is assigned a time on that method, or a time on no method, as relevant. These times are based on accelerated failure time models using the contraceptive calendar data from the 2022 Kenya DHS. In this case, we use the 2022 Kenya DHS to increase our sample size. Method-specific failure times are more computationally intensive to calculate than simple annual probabilities, and we determined a need for a larger sample size to allow the models to converge.

For each woman in the data, we identified the first time she switched a method from one month to the next. From when she switched to that method, we then counted how many months she continued to use that method, up until the end of the contraceptive calendar. We then fit accelerated failure time models for each method and compared several distributions: exponential, Weibull, log-logistic, log-normal, Gompertz, and gamma. The models included age groups as a factor to understand its impact on the time to discontinuation. Model fit was assessed by plotting fitted survival curves against the empirical survival data and selecting the distribution with the lowest AIC.

We then produced coefficients and 95% pointwise confidence bands from the model with the best fitting distribution for each method and implemented them as parameters in FPsim v2.0. These parameters are used to produce a probability distribution function (Figure 4) to select an individualized time on method (or no method).

**Figure 4.**
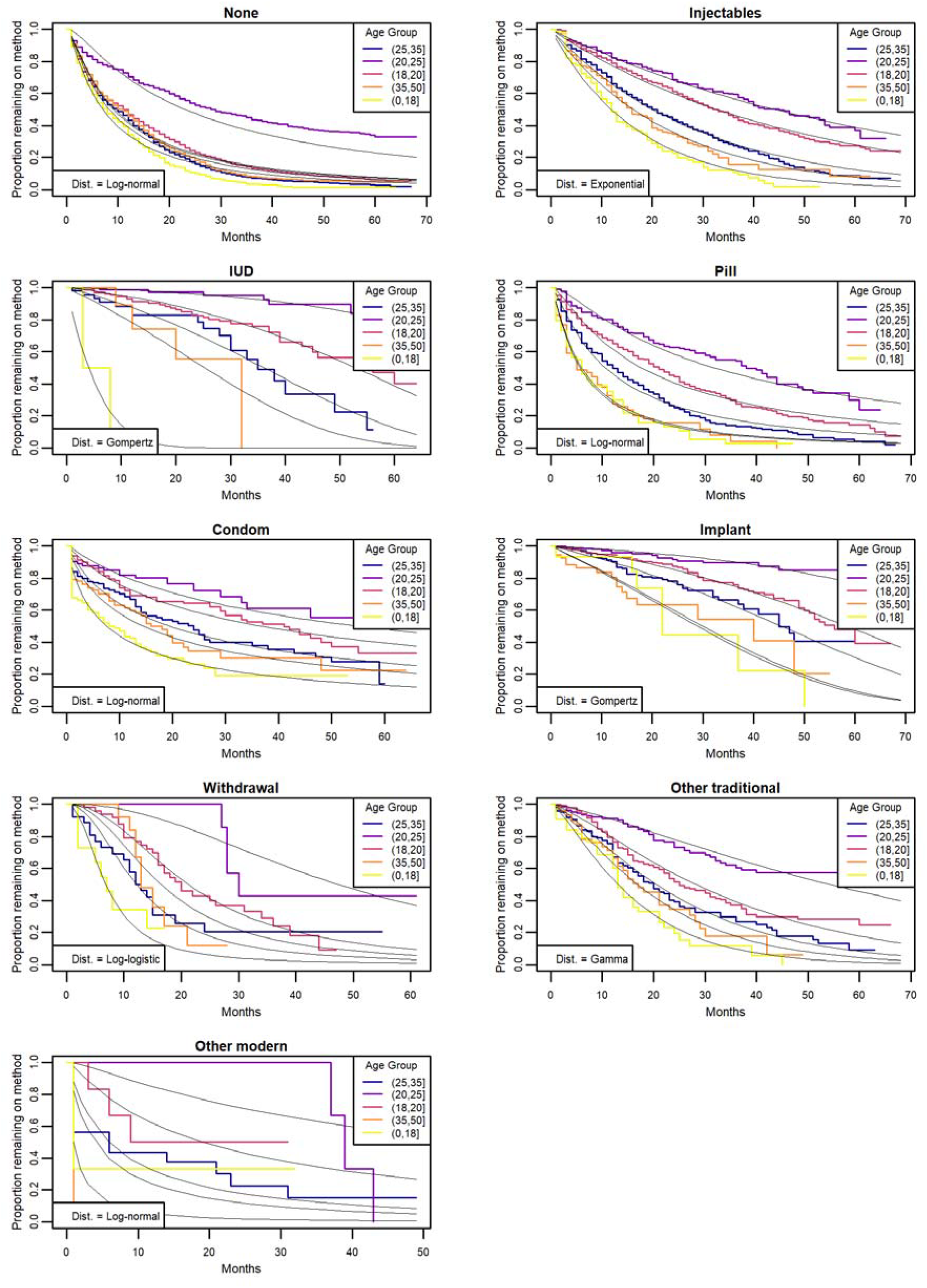
Fitted accelerated failure time models for expected time on each contraceptive method. Each panel is a contraceptive method, colors are the Kaplan-Meier curves for each method, and black lines are the fitted model with the distribution listed on the bottom left, which was selected due to lowest AIC.

**Figure 5.**
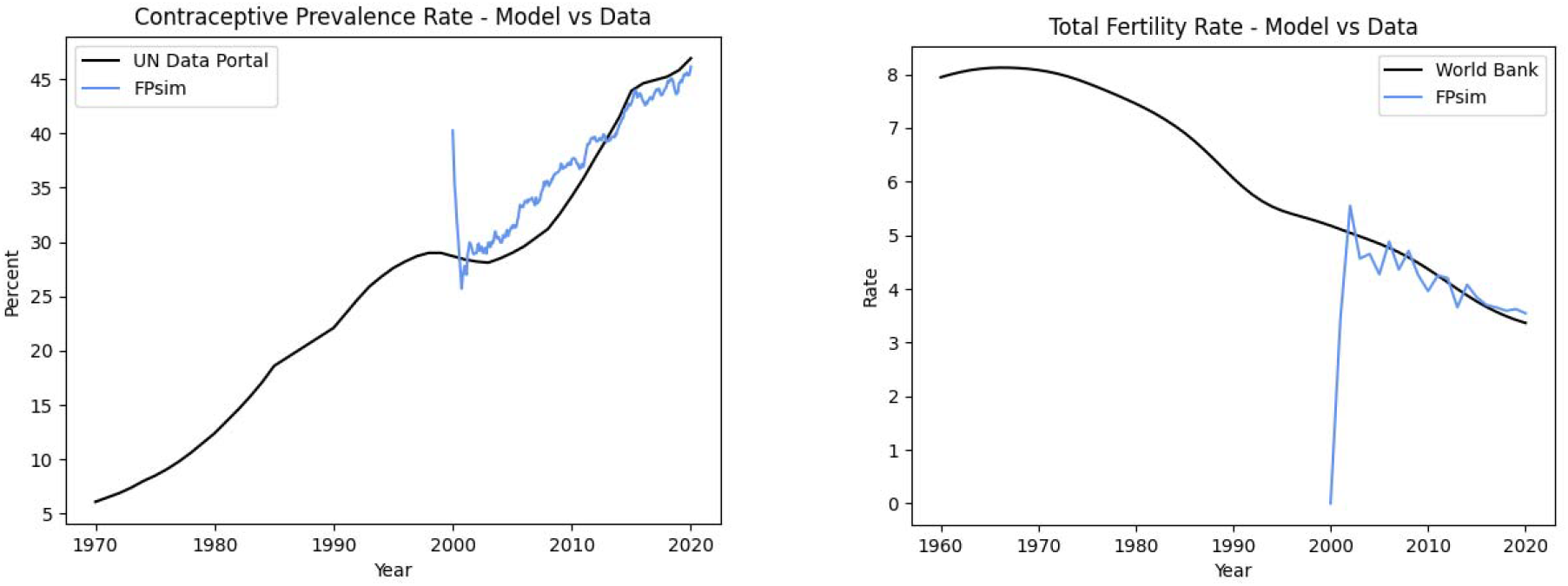
Plots of CPR (contraceptive prevalence rate) and TFR (total fertility rate) calibration targets over time. Black lines represent empirical data sources—UN Data Portal estimates for CPR (top) and World Bank estimates for TFR (bottom). Blue lines show FPsim 2.0 simulation outputs from the Detailed Choice model, calibrated to a Kenya-like setting. These plots highlight the model’s ability to reproduce historical trends and project future trajectories in CPR and TFR, demonstrating strong alignment with observed data from the mid-2000s onward.

### Calibration

Calibration of agent-based models to data is a crucial step to produce results that inform public health policy(18). FPsim users can perform both manual and automatic calibration with the option to weigh the calibration targets that are most important for their work. The automatic calibration process takes in any parameter and optimizes it to reduce mismatch between a given simulation and data provided. Most commonly, FPsim calibrations aim to reduce mismatch for age-specific fertility rates, contraceptive prevalence, and contraceptive method mix.

Because the model incorporates many free parameters that rely on theoretical assumptions where data are scarce, the process of calibration often involves adjustment of these free parameters within a plausible bound. For instance, biological fecundability is an age-specific pattern in FPsim, but we also recognize that individual heterogeneity is a critical component in understanding fertility in a population. Because we do not have enough data on individual-level heterogeneity, nor do we fully understand the underlying mechanisms that produce more or less fecundability for individuals beyond genetic makeup, we use free parameters to adjust fecundability for each individual based on a random multiplier. Thus, some individuals will always have higher than average fecundability for their age, and some individuals will always have lower fecundability for their age. The upper and lower bounds on this individual heterogeneity are free parameters, which can be adjusted during the calibration process.

The FPsim v2.0 calibration to a Kenya-like setting includes key fertility and family planning calibration targets. We used an iterative calibration process for the FPsim v2.0 model, beginning with the least complex contraceptive choice model, “Simple Choice,” before progressing to the more complex “Detailed Choice.” The detailed choice module introduced additional covariates and refinements to align the model with observed trends in contraceptive prevalence rate (CPR), total fertility rate (TFR), method mix, age-specific fertility rate (ASFR), age at first birth, and birth spacing. These parameters served as primary calibration targets.

To ensure a robust assessment of each model calibration, we implemented a systematic approach that accounts for both scale differences across calibration targets and variability in model outputs. Specifically, we conducted 30 independent model runs using different random seeds to capture stochastic variation in the simulation. For each calibration target—Age-Specific Fertility Rate (ASFR), Total Fertility Rate (TFR), Contraceptive Prevalence Rate (CPR), Method Mix, Age at First Birth (AFB), and Birth Spacing, we applied Z-score normalization to both the model oututs and empirical data. This step was essential to standardize metrics with different scales, ensuring that no single target disproportionately influenced the overall calibration assessment.

Following normalization, we computed the Mean Squared Error (MSE) between the model and observed data for each calibration target across the 30 model runs. We then averaged these MSE values per calibration target to obtain a representative measure of model performance for each metric. Finally, we computed the mean and standard deviation of the MSEs across all calibration targets, yielding an overall accuracy score. This approach enables a comprehensive evaluation of model fit, balancing the contributions of multiple demographic and behavioral indicators while quantifying uncertainty in the model’s predictive performance.

### Calibration of the “Simple Choice” Model

Adjustments and enhancements during this phase of calibration included:

1. **Refinement of free parameters:** Calibration focused on adjusting free parameters to best fit the data (see Table 1 for specific parameters used).
2. **Incorporation of a CPR time trend term:** The parameters *prob_use_year, prob_use_trend_par*, and *prob_use_intercept* (see Table 1) were incorporated into the logistic function as the base year, trend value, and offset parameter to use for probability of contraceptive use. The function was redefined as:

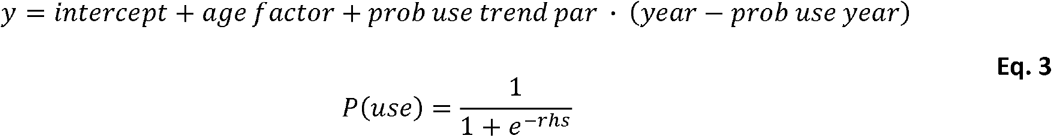

These terms were also added as contraceptive module free parameters and were instrumental in calibrating the model by fine-tuning CPR trends.
3. **Addition of ‘*ever_used’* (contraception) as a predictor for probability of using contraception:** The initial version of the “simple choice” model did not include previous use of contraception in the probability calculation. It was added at this phase of model development. This addition helped address the underrepresentation of implants and injectables by increasing the likelihood of long-acting method use among women with prior contraceptive use.
4. **Incorporation of method weights:** Method weights were introduced as free parameters to scale the probabilities of choosing each contraceptive method. These weights were re-normalized to ensure they did not directly translate into disproportionate likelihoods.

**Table 1.** Free parameters used to iteratively calibrate the model.

| Free Parameter | Definition | Models Calibrated |
| --- | --- | --- |
| *fecundity_var_low* | used to stretch fecundity by a factor bounded by [fecundity_var_low, fecundity_var_high] | Simple, Detailed |
| *fecundity_var_high* | used to stretch fecundity by a factor bounded by [fecundity_var_low, fecundity_var_high] | Simple, Detailed |
| *exposure_factor* | overall scale factor on the probability of becoming pregnant | Simple, Detailed |
| *prob_use_year* | A reference year used as the baseline for calculating the trend in contraceptive use probability | Simple, Detailed |
| *prob_use_trend_par* | A scaling factor that determines the strength and direction of the time trend in contraceptive use probability. It defines how much the probability changes per unit year relative to *prob_use_year* | Simple, Detailed |
| *method_weights* | A set of scaling factors applied to the initial probability distribution of contraceptive method uptake, adjusting the likelihood of selecting each method | Simple, Detailed |
| *exposure_correction_age* | An array of factors to be applied to account for residual exposure to either pregnancy or live birth by age | Detailed |
| *exposure_correction_parity* | An array of factors to be applied to account for residual exposure to either pregnancy or live birth by parity | Simple, Detailed |

To identify discrepancies in contraceptive prevalence rate (CPR) trends and refine model calibration, we used age-stratified plots of CPR and method mix, which allowed for a detailed comparison of modeled versus observed contraceptive use patterns across different age groups. Additionally, comparing contraceptive access to new users was instrumental in diagnosing potential mismatches between the supply of contraception and uptake dynamics. These visual diagnostics helped pinpoint specific areas where the model deviated from empirical data, guiding parameter adjustments to improve overall alignment.

### Calibration of the “Detailed Choice” Model

The initial free parameters and enhancements were carried over from the final “Simple Choice” calibration described above. Similar figures were used for visualization and troubleshooting, with the addition of covariate plots. Adjustments and enhancements during this phase of calibration included:

1. **Refinement of free parameters:** Calibration focused on adjusting free parameters to best fit the data (see Table 1 for specific parameters used). This included the modification of the *exposure_correction_age* free parameter, which accounted for residual pregnancy and live birth exposure by age. This adjustment greatly improved the alignment of age-specific fertility rates (ASFR) with observed data, particularly by addressing the elevated rates in older age groups, which could not be resolved through adjustments to *exposure_correction_parity* alone.
2. **Addition of interaction term between age and ever use of contraception:** This interaction proved essential for improving CPR results, producing reasonable trends after a 5-year burn-in period.

## Results

### Calibration Performance

The calibration of the “Simple Choice” model resulted in the following measurement, based on the targets aforementioned: Overall MSE Score: **0.2927 ± 0.5138**, with the following MSE values per target:

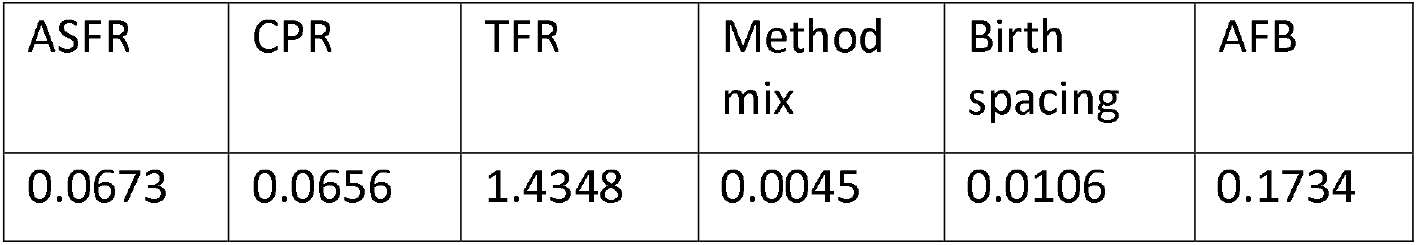

| ASFR | CPR | TFR | Method mix | Birth spacing | AFB |
| --- | --- | --- | --- | --- | --- |
| 0.0673 | 0.0656 | 1.4348 | 0.0045 | 0.0106 | 0.1734 |

The calibration of the “Detailed Choice” model resulted in the following measurement, based on the targets aforementioned: Overall MSE Score: **0.0868 ± 0.0625**, with the following MSE values per target:

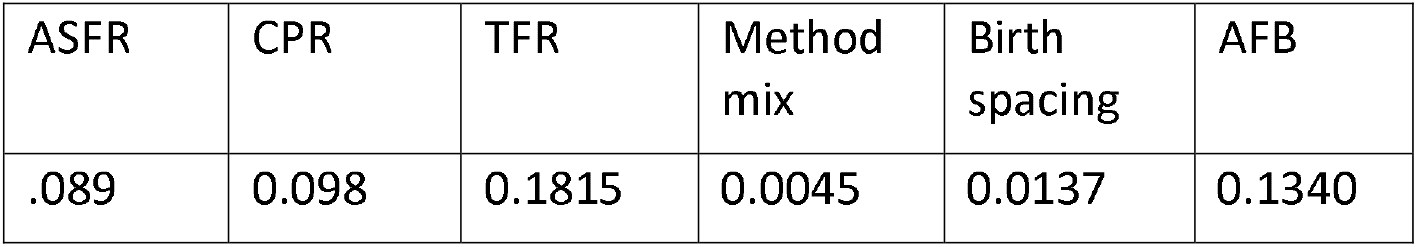

### Model Fit and Utility

The model results in Figure 6 demonstrate a strong overall calibration across key demographic and reproductive health indicators, with an overall MSE score of 0.0868 ± 0.0625 for the Detailed Choice model, indicating a close alignment with observed data.

**Figure 6.**
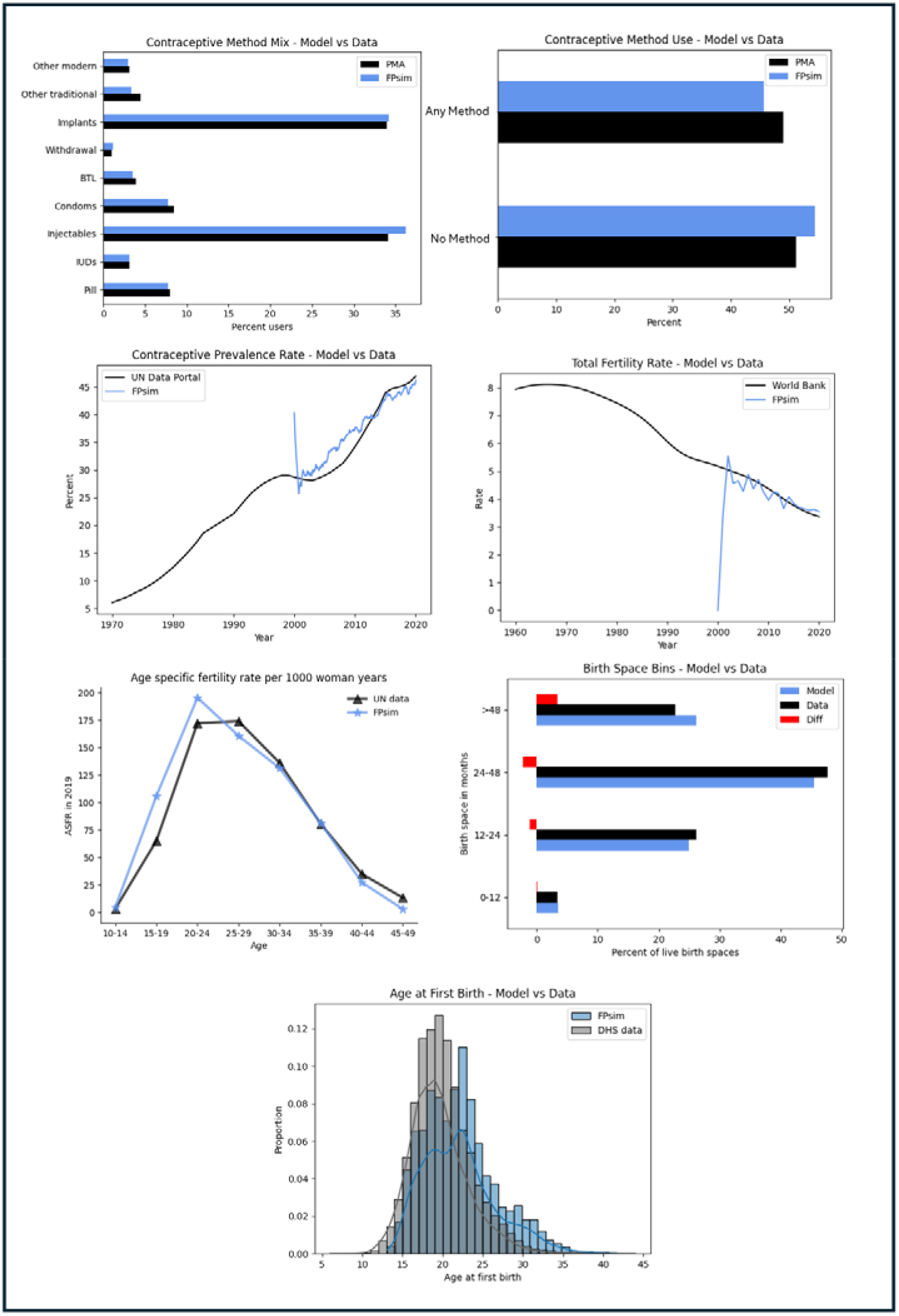
Calibration results for FPsim 2.0 using the Detailed Choice model in a Kenya-like setting. The model was calibrated to multiple demographic and reproductive health indicators, including contraceptive use, fertility, and birth ages/spacing. Each panel compares FPsim outputs (blue) against empirical data (black or gray) from sources such as PMA, DHS, UN, and the World Bank. The panels show model alignment with method mix, method use, contraceptive prevalence rate (CPR), total fertility rate (TFR), age-specific fertility rates (ASFR), birth spacing distributions, and age at first birth. Collectively, these outputs demonstrate strong model fit across key behavioral and demographic indicators, supporting the model’s validity for simulating complex family planning and empowerment dynamics in the Kenyan context.

Age-Specific Fertility Rate (ASFR) (MSE = 0.089) and Contraceptive Prevalence Rate (CPR) (MSE = 0.098) are well captured, reflecting the model’s ability to accurately reproduce fertility trends and contraceptive use patterns. Similarly, the contraceptive method mix shows an exceptionally low MSE (0.0045), suggesting precise modeling of contraceptive preferences. Birth spacing (MSE = 0.0137) and age at first birth (MSE = 0.134) exhibit strong agreement with empirical data, reinforcing the model’s robustness in capturing reproductive behavior.

Overall, the calibration results indicate that the model effectively reproduces key demographic and behavioral trends. The strong fit across most targets highlights the model’s reliability in simulating broader population dynamics.

## Discussion

### Advancing Person-Centered Family Planning Modeling

In this study, we significantly enhanced FPsim, an agent-based model for FP, by incorporating additional demographic metrics such as educational attainment, wealth, and urbanity. These improvements aimed to provide a more nuanced, person-centered understanding of FP behaviors and outcomes. We demonstrated the feasibility and utility of these enhancements through an application of the model to a Kenya-like setting, leveraging data from the Kenya PMA surveys.

### Methodological Innovations

The methodological enhancements presented in this paper contribute to the field of agent-based modeling in several ways. First, by moving from an annual probability of using contraception framework to a multifactorial, time-specific probability model, we align FPsim v2.0 more closely with real-world contraceptive use patterns. This approach allows for more precise estimation of method duration and switching behaviors, which are crucial for understanding the effectiveness and satisfaction with different contraceptive methods. Second, the implementation of individualized time-on-method distributions based on empirical data represents a methodological innovation that improves the model’s realism and predictive accuracy. This refinement acknowledges the diverse experiences of women using different contraceptive methods and allows for more tailored policy recommendations. Lastly, the incorporation of individual-level demographic attributes—such as education, wealth, and urbanicity—substantially improved the model’s ability to replicate observed patterns in contraceptive use and fertility. These features enable the model to more accurately reflect the heterogeneity of real-world experiences and behaviors, particularly in settings marked by social and economic disparities.

### Policy and Programmatic Implications

The practical implications of these model enhancements are substantial. By providing a more detailed and accurate simulation of FP behaviors, FPsim v2.0 can inform more effective and context-specific FP policies and programs. For instance, understanding the factors that influence contraceptive discontinuation can help design interventions that address these barriers, ultimately reducing unintended pregnancies and improving reproductive health outcomes. Policymakers can use FPsim v2.0 to test hypothetical scenarios and identify strategies that not only increase contraceptive prevalence but also enhance women’s decision-making autonomy and economic opportunities. In that way, FPsim v2.0 can be used to demonstrate the broader social and economic impacts of family planning programming.

### Model Limitations and Future Directions

Despite these advancements, several limitations must be acknowledged. The reliance on PMA data, while robust, may not capture all relevant factors influencing FP behaviors. Future iterations of FPsim could benefit from integrating additional data sources and exploring other dimensions not included in the current model. Additionally, while the model’s calibration to a Kenya-like setting demonstrates its applicability, further validation and calibration in other contexts are necessary to ensure its generalizability. Expanding the model’s applicability to other countries and regions with distinct cultural, socioeconomic, and policy environments will enhance its utility and relevance.

### Insights and Learnings from the Calibration Process

The iterative calibration process highlighted how systematically incorporating demographic detail improved model fit and interpretability. Starting with simpler modules—like the “Simple Choice” model—enabled us to isolate and resolve structural misalignments in key outcomes such as contraceptive prevalence rate (CPR) and age-specific fertility rates (ASFR). Once baseline alignment was achieved, layering in additional covariates like wealth quintile, urban residence, and educational attainment consistently improved alignment with observed subgroup trends, confirming their critical role in shaping reproductive behavior.

Additionally, the calibration process highlighted the importance of free parameters in capturing heterogeneity in biological and behavioral factors. The inclusion of free parameters to adjust fecundability and exposure was crucial in aligning contraceptive prevalence rate (CPR) and age-specific fertility rates (ASFR) with observed trends. The use of free parameters like *exposure_correction_parity* and *exposure_correction_age* further emphasized the need to account for age- and parity-specific variations in reproductive behavior, particularly in addressing discrepancies in older age groups.

Integrating covariate-specific plots for troubleshooting (e.g., parity, urban residence, and educational attainment over time) was instrumental in diagnosing trends and refining parameterization. This iterative visualization approach not only facilitated calibration but also improved transparency and model interpretability.

Overall, the iterative calibration process allowed for a gradual incorporation of complexity while ensuring robust alignment with empirical data. This approach highlighted the value of modular calibration, visualization, and targeted parameter adjustments in building a reliable and interpretable agent-based model for family planning.

## Conclusions

The enhancements introduced in FPsim v2.0 represent a significant step toward a more holistic and person-centered approach to modeling family planning behaviors and outcomes. By incorporating additional dimensions of social and economic context – such as education, wealth, and urban residence – into the simulation framework as well as refining the estimation of method duration and switching behaviors, FPsim now captures the diversity of real-world contraceptive experiences with greater precision and nuance. These improvements not only increase the model’s empirical fidelity but also expand its relevance for addressing equity and access in reproductive health. As a result, FPsim v2.0 offers a robust and flexible tool for researchers, policymakers, and program designers seeking to understand and address the complex drivers of contraceptive use and discontinuation. As we continue to refine FPsim, it holds great potential to inform more effective, equitable, and context-specific family planning strategies worldwide.

## Data Availability

All data referenced in this paper are publicly available.

https://dhsprogram.com/data/

https://www.pmadata.org/data

## Footnotes

1 Code, documentation, and tutorials are openly available at fpsim.org

